# Assessment of Erectile Dysfunction in Patients with Benign Prostate Hyperplasia Using the Sexual Function Index at Muhimbili National Hospital, Dar es Salaam, Tanzania

**DOI:** 10.1101/2023.03.06.23286860

**Authors:** Said Ali Mawji, Charles Mkony, Obadia Nyongole, Peter Msinde, Deogratius Mahenda

**Affiliations:** School of Surgery, Muhimbili University of Health and Allied Sciences, Dar es Salaam-Tanzania; Urology Department, Muhimbili National Hospital, Dar es Salaam-Tanzania

**Keywords:** Benign prostatic enlargement, erectile dysfunction, IIEF tool

## Abstract

**Background:** Erectile dysfunction (ED) is the inability to achieve and maintain a steady erection for sexual performance. Erectile dysfunction (ED) is becoming more common in the general population, particularly among elderly men with lower urinary tract symptoms and those with benign prostatic hyperplasia. This rise in erectile dysfunction has also been attributed by several other etiologies, which includes inflammatory, mechanical, psychological, occlusive vascular, trauma, endocrine, neurologic, chemical and extra factors (prostatectomy, old age). The purpose of the study was to assess erectile dysfunction in patients with benign prostate hyperplasia before treatment using international index of erectile function (IIEF) tool.

**Methodology:** Hospital-based cross-sectional study from March 2021 to June 2022, which involved 188 patients clinically diagnosed with benign prostatic enlargement. Convenient sampling method was used to recruit participants and structured questionnaires were used to attain social demographic characteristics. The international prostate symptom score (IPSS) was used to assess, diagnose patients with BPH as well as the severity of LUTS and international index of erectile function (IIEF) was used to assess erectile dysfunction. Data was analyzed using SPSS software version 27.

**Results:** The proportion of ED in patients with benign prostate hyperplasia/LUTS using IIEF tool was found to be 154(82%) and those with no erectile dysfunction was 34(18%). The factors that were statistically associated with erectile dysfunction were age, marital status, cigarette smoking, and diabetes mellitus.

**Conclusion:** Erectile dysfunction is very common in men who have Benign Prostate Hyperplasia. The prevalence and severity of ED increased with age, diabetes, cigarette smoking, lower urinary tract symptoms, and hypertension, according to the study.

## Background

Erectile dysfunction (ED) is commonly described as the inability to achieve or maintain an erection that is satisfactory for the completion of sexual intercourse.

The excessive cellular proliferation of the glandular and stromal parts of the prostate is known as benign prostatic hyperplasia (BPH). It’s one of the most frequent conditions affecting adult men, with the frequency rising drastically after 50. BPH symptoms, such as difficulty peeing, urgency with leaking or dribbling, and nocturia, can impair sexual functioning and other elements of quality of life (QOL), but therapy can also impair QOL by causing sexual dysfunction. As a result, many men with BPH and lower urinary tract symptoms (LUTS) put off seeking medical help until their symptoms become unbearable. Sexual problems, including as erectile dysfunction (ED) and ejaculatory problems, are usually linked to BPH-related LUTS. Interest in sexual intercourse reduces as the severity of LUTS increases. Men with severe LUTS have lower libido, have a harder time maintaining an erection, and have less sexual satisfaction than men with mild LUTS. (1)

LUTS symptoms are classified as storage (increased daytime frequency, nocturia, urgency, and/or urinary incontinence), voiding (low/weak stream, terminal dribble, and/or hesitancy), and post micturition (incomplete emptying and post-micturition dribble), and can also include sexual intercourse and genital and lower urinary tract pain. Overactive bladder (OAB) symptoms might include urgency with or without urgent incontinence, whereas voiding issues in men can indicate BOO and/or BPH (2). Lower urinary tract symptoms (LUTS) caused by benign prostatic hyperplasia have been associated to sexual dysfunction. Erectile dysfunction (ED) and ejaculatory dysfunction are two types of sexual dysfunction (premature ejaculation, retrograde ejaculation, delayed ejaculation, painful ejaculation, and decreased ejaculation force). (3)

IPSS is a questionnaire for assessing BPH that is self-administered by the patient. a screening tool that can be used to quickly diagnose, track, and propose treatment for BPH symptoms. It includes seven questions about BPH symptoms as well as one question about the patient’s subjective quality of life. Each of the first seven questions relates to urinary symptoms, and the patient is given the option of selecting one of six answers reflecting the severity of the condition. The answers are graded on a scale of 0 to 5. As a result, the total score can range from 0 to 35. (asymptomatic to very symptomatic). All of the questions are about the seven urinary symptoms listed below: (Frequency, Intermittency, Urgency, Weak Stream, Straining, Nocturia, Incomplete emptying) Mild (symptom score 7), Moderate (symptom score 8–19), and Severe (symptom score 20–35) are the three levels of symptom severity. Even though IPSS was originally validated in males with BPH, it is also applicable to women. When compared to younger individuals, elderly people have a much higher IPSS score. The severity of erectile dysfunction is linked to the IPSS score’s symptoms. The ED is worsened by the severity of LUTS/BPH symptoms and a higher IPSS score. The IIEF score is inversely linked to the symptoms of the IPSS score, with a rise in the IPSS level resulting in a decrease in the IIEF score. (20)

Organic ED is frequently linked to chronic illnesses like heart disease and diabetes. It’s a sign of vascular endothelium injury, which can be caused by hypertension, diabetes, high cholesterol, or smoking. (4)

LUTS suggestive of BPH affect a large percentage of older men, and the condition’s prevalence increases with age. Similarly, as people get older, the prevalence of sexual dysfunction increases. The most common symptoms of sexual dysfunction include erectile dysfunction (ED), ejaculatory irregularities, low libido (also known as hypoactive sexual desire), or a combination of these symptoms. While it was often thought that LUTS/BPH and sexual dysfunction were just co-existing illnesses in elderly men, new research shows that sexual dysfunction is significantly more common in LUTS/BPH patients than in men without the disease, even when age and comorbidity are taken into account. As a result, it was determined that LUTS/BPH is a separate component in the development of sexual dysfunction. This link has prompted the question of whether LUTS/BPH and sexual dysfunction share an underlying disease. (7). In clinical trials, several self-administered questionnaires have been developed to assess sexual functioning. They give important data on the prevalence and severity of sexual symptoms in community and clinic populations, as well as therapeutic implications. (8). The IIEF Questionnaire was created to assess the influence of erectile dysfunction on a person’s reproductive health during the previous four weeks. Finally, results were divided into five categories by the 15-point questionnaire: erectile function, orgasmic function, sexual desire, intercourse satisfaction, and overall satisfaction. “No malfunction” to “severe dysfunction” were the clinical interpretations. (9).

The goal of this study is to use the IIEF Questionnaire, a validated instrument, to assess patients with benign prostatic hyperplasia/LUTS experienced erectile dysfunction before starting treatment.

## Methods

### Study design and setting

The study was a prospective cross-sectional study design conducted in Dar es Salaam, Tanzania at Muhimbili National Hospital (MNH).

### Study population

The study population consisted of all eligible male patients above 40 years old with lower urinary tract symptoms/Benign Prostatic Enlargement (BPE) which was identified by clinical examination at MNH attending Urology clinics and Urology inpatients wards March 2021 to June 2022.

### Sample size

Sample size was calculated using the Yamane’s (1967) methodology to calculate the minimum required sample size as stated below:

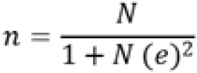

Where n denotes the needed minimum sample size and N is the number of participants. (all eligible male patient where the minimum number per month is 50) e is the precision level, which is set at 5%. (the precision level given a 95 percent power of the study). The degree of confidence is 95%.

Therefore;

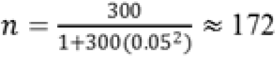

A 10% will be added for the non-responses, thus

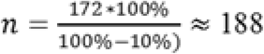

The minimum sample size of will be 188 eligible male patients.

### Sampling method

Convenience sampling was used to recruit the study participants from MNH Urology clinics and Urology inpatients until the required sample size was reached. Patients were screened for inclusion and exclusion criteria in order to identify those who were eligible to participate in the study.

### Data collection methods

The purpose of the study was explained to eligible patients, and they were requested to consent to participate. Questionnaires were given by the trained assistants who are trained doctors working in the Urology clinics and inpatient wards.

Each questionnaire evaluated Erectile Dysfunction based on the International Index of Erectile Function. The IIEF is a validated questionnaire consisting of 15 questions, all the questions break down into five specific areas, which are Erectile function (Qn 1-5 & 15) with maximum score of 30, Orgasmic function (Qn 9-10) with maximum score of 10, Sexual desire (Qn 11-12) with maximum score of 10, Intercourse satisfaction (Qn 6-8) with maximum score of 15, and Overall satisfaction (Qn 13-14) with maximum score of 10. Each scored from 0 to 5, in which a global score below 21 is considered significant for ED, scores were classified as 21-25 to be normal Erectile function,16- 20 to be mild Erectile Dysfunction, 11-15 points to be moderate ED, and less than 10 to be severe ED. The questionnaire also included information on socio demographic information, diabetes mellitus, cigarette smoking, hypertension, high cholesterol, prostate disease, ischemic heart disease, pelvic injuries and lower urinary tract symptoms using IPSS chart were used.

### Data management and analysis

The data was captured in the questionnaires that were grouped and examined for errors using Epi info version 7.2, then the data cleaning procedure was conducted. The processed data was analyzed using Statistical Package for the Social Sciences (SPSS) version 27 software. Descriptive analysis of sociodemographic characteristics was done using frequencies, means and percentages.

The categorical variables were expressed in percentages and the association between dependent and independent variables were analyzed using bivariate and multivariate linear regression models, the dependent variable being erectile dysfunction from the questionnaire. Co-efficient intervals were considered a measure of association, with a p-value of less than 0.05 considered for statistical significance. Statistically significant independent variables were then compared to measure the strength of association using Pearson’s correlation coefficients.

## Results

### Socio-demographic characteristics of the study participants

The study involved 188 participants. The mean age was 71 with range of 50 to 98 years. All participants in this study had Benign Prostate Enlargement.

The study revealed that the proportion of ED in patients with benign prostate hyperplasia/LUTS using IIEF tool was 154(82%) and those with no erectile dysfunction was 34(18%)

**Figure 2:**
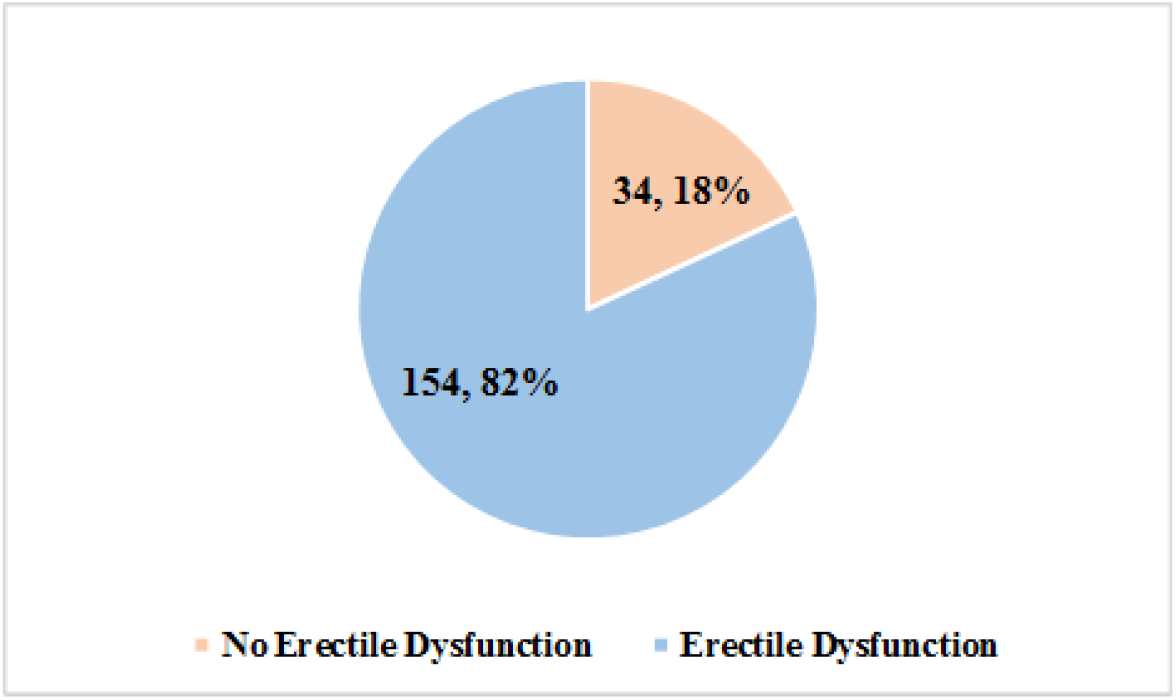
The pie chart showing the proportion of ED in patients with BPH/LUTS using IIEF tool.

### The factors associated with erectile dysfunction presented by patients with benign prostate hyperplasia

As shown in table 2 those participants with 71 years and above had 92.94% erectile dysfunction. The age was statistically significantly associated with erectile dysfunction among the participants (p = <0.001).

**Table 1:** Social demographic characteristics among the participants.

| **Variable** | **Frequency** | **Percent (%)** |
| --- | --- | --- |
| **Education category** |  |  |
| No formal | 70 | 37.23 |
| Primary | 51 | 27.13 |
| secondary | 43 | 22.87 |
| collage/ university | 24 | 12.77 |
| **Occupation category** |  |  |
| Employed | 114 | 60.64 |
| Unemployed | 74 | 39.36 |
| **Marital status** |  |  |
| Married | 136 | 72.34 |
| Not married | 32 | 17.02 |
| Co-habiting | 20 | 10.64 |
| **Age group** |  |  |
| <61 | 36 | 19.15 |
| 61 to 70 | 67 | 35.64 |
| ≥71 | 85 | 45.21 |
| **Cigarette Smoking** |  |  |
| Yes | 62 | 32.98 |
| No | 126 | 67.02 |
| **IPSS score category** |  |  |
| Mild | 38 | 20.21 |
| Moderate | 74 | 39.36 |
| Severe | 76 | 40.43 |
| **Experiencing loss libido** |  |  |
| Yes | 26 | 13.83 |
| No | 162 | 86.17 |

**Table 2:** The factors associated with erectile dysfunction presented with BPH patients.

| **Variable** | **Negative**<br>**Freq (%)** | **Positive**<br>**Freq (%)** | **Total**<br>**N** | **P value** |
| --- | --- | --- | --- | --- |
| **Age Group** |  |  |  |  |
| Less 61 | 16(44.44) | 20(55.56) | 36 | **<0.001** |
| 61 to 70 | 12(17.91) | 55(82.09) | 67 |  |
| ≥71 | 6(7.06) | 79(92.94) | 85 |  |
| **Education Level** |  |  |  |  |
| No formal | 11(15.71) | 59(84.29) | 70 | 0.055 |
| Primary | 9(17.65) | 42(82.35) | 51 |  |
| Secondary | 5(11.63) | 38(88.37) | 43 |  |
| Collage/ university | 9(37.5) | 15(62.5) | 24 |  |
| **Occupation category** |  |  |  |  |
| Employed | 10(8.77) | 104(91.23) | 114 | **<0.001** |
| Unemployed | 24(32.43) | 50(67.57) | 74 |  |
| **Marital status** |  |  |  |  |
| Married | 21(15.44) | 115(84.56) | 136 | 0.105 |
| Not married | 6(18.75) | 26(81.25) | 32 |  |
| Co-habituating | 7(35) | 13(65) | 20 |  |
| **Cigarette Smoking** |  |  |  |  |
| Yes | 6(9.68) | 56(90.32) | 62 | **0.036** |
| No | 28(22.22) | 98(77.78) | 126 |  |
| **Diabetes status** |  |  |  |  |
| Yes | 1(3.57) | 27(96.43) | 28 | **0.032\*** |
| No | 33(20.63) | 127(79.38) | 160 |  |
| **HBP-Category score category** |  |  |  |  |
| Yes | 4(19.81) | 33(89.19) | 37 | 0.241* |
| No | 30(19.87) | 121(80.13) | 151 |  |
| **High cholesterol** |  |  |  |  |
| Yes | 5(23.81) | 16(76.19) | 21 | 0.470 |
| No | 29(17.37) | 138(82.63) | 167 |  |
| **Ischemic heart disease** |  |  |  |  |
| Yes | 9(13.04) | 60(86.96) | 69 | 0.238 |
| No | 25(21.01) | 94(78.99) | 119 |  |
| **Pelvic injuries** |  |  |  |  |
| Yes | 4(17.39) | 19(82.61) | 23 | 1.00* |
| No | 30(18.18) | 135(81.82) | 165 |  |
| **IPSS Score Category** |  |  |  |  |
| Mild | 15(39.47) | 23(60.53) | 38 | **<0.001** |
| Moderate | 18(24.32) | 56(75.68) | 74 |  |
| Severe | 1(1.32) | 75(98.68) | 76 |  |
| **Experiencing loss libido** |  |  |  |  |
| Yes | 6(18.75) | 26(81.25) | 32 | 0.915 |
| No | 28(17.95) | 128(82.05) | 156 |  |

**Table 3:**
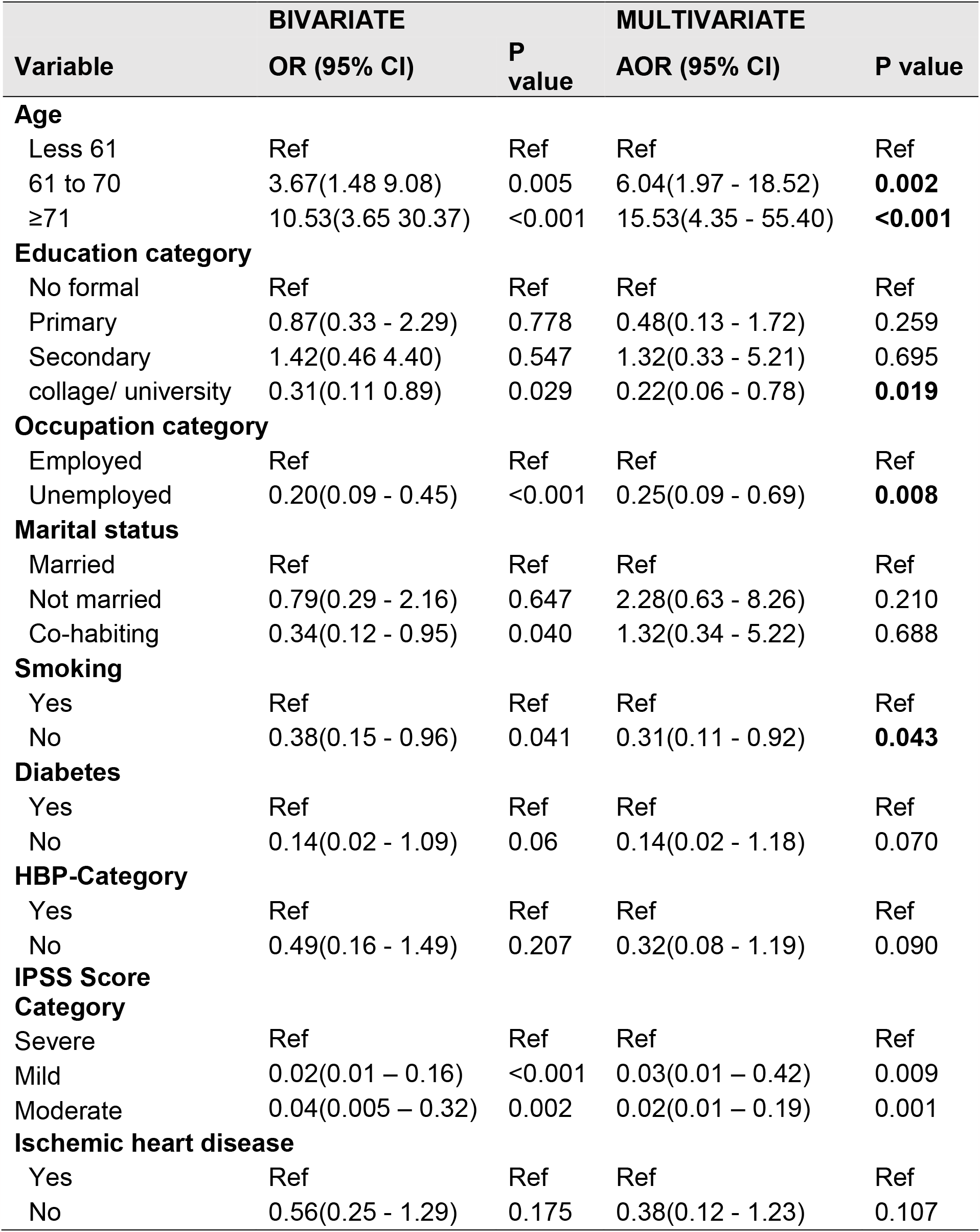
The factors associated with BPH using bivariate and multivariate analysis by binary logistic regression model.

| Variable | BIVARIATE<br>OR (95% CI) | P<br>value | MULTIVARIATE<br>AOR (95% CI) | P value |
| --- | --- | --- | --- | --- |
| **Age** |  |  |  |  |
| Less 61 | Ref | Ref | Ref | Ref |
| 61 to 70 | 3.67(1.48 9.08) | 0.005 | 6.04(1.97 - 18.52) | **0.002** |
| ≥71 | 10.53(3.65 30.37) | <0.001 | 15.53(4.35 - 55.40) | **<0.001** |
| **Education category** |  |  |  |  |
| No formal | Ref | Ref | Ref | Ref |
| Primary | 0.87(0.33 - 2.29) | 0.778 | 0.48(0.13 - 1.72) | 0.259 |
| Secondary | 1.42(0.46 4.40) | 0.547 | 1.32(0.33 - 5.21) | 0.695 |
| collage/ university | 0.31(0.11 0.89) | 0.029 | 0.22(0.06 - 0.78) | **0.019** |
| **Occupation category** |  |  |  |  |
| Employed | Ref | Ref | Ref | Ref |
| Unemployed | 0.20(0.09 - 0.45) | <0.001 | 0.25(0.09 - 0.69) | **0.008** |
| **Marital status** |  |  |  |  |
| Married | Ref | Ref | Ref | Ref |
| Not married | 0.79(0.29 - 2.16) | 0.647 | 2.28(0.63 - 8.26) | 0.210 |
| Co-habiting | 0.34(0.12 - 0.95) | 0.040 | 1.32(0.34 - 5.22) | 0.688 |
| **Smoking** |  |  |  |  |
| Yes | Ref | Ref | Ref | Ref |
| No | 0.38(0.15 - 0.96) | 0.041 | 0.31(0.11 - 0.92) | **0.043** |
| **Diabetes** |  |  |  |  |
| Yes | Ref | Ref | Ref | Ref |
| No | 0.14(0.02 - 1.09) | 0.06 | 0.14(0.02 - 1.18) | 0.070 |
| **HBP-Category** |  |  |  |  |
| Yes | Ref | Ref | Ref | Ref |
| No | 0.49(0.16 - 1.49) | 0.207 | 0.32(0.08 - 1.19) | 0.090 |
| **IPSS Score Category** |  |  |  |  |
| Severe | Ref | Ref | Ref | Ref |
| Mild | 0.02(0.01 – 0.16) | <0.001 | 0.03(0.01 – 0.42) | 0.009 |
| Moderate | 0.04(0.005 – 0.32) | 0.002 | 0.02(0.01 – 0.19) | 0.001 |
| **Ischemic heart disease** |  |  |  |  |
| Yes | Ref | Ref | Ref | Ref |
| No | 0.56(0.25 - 1.29) | 0.175 | 0.38(0.12 - 1.23) | 0.107 |

Also, the study revealed that participants who were past smokers or current smokers had statistically significant association with erectile dysfunction (p=0.036). Furthermore, the study found that participants who had diabetes mellitus had statistically significant association with erectile dysfunction, since (P=0.032).

### Bivariate and multivariate model using logistic regression to assess the factors associated with Erectile dysfunction

Participants with age group between 61 and 70 had the odds ratio 6.04 (95% CI: 1.97 - 18.52) times more likely to have ED, which is statistically significant (p=0.002). The participants with age group 71 or greater had the odds ratio 15.53(95% CI: 4.35 - 55.40) times more likely to have ED, which is statistically significant (p=0.001). The participants with college or university education level had the odds ratio 0.22(95% CI: 0.06 - 0.78) times less likely to have ED compared with those who had no formal education level which is statistically significant (p=0.019). The participants who were unemployed had the odds ratio 0.25(95% CI: 0.09 - 0.69) times less likely compared with those employed which is statistically significant (p=0.008). The participants who were current or past nonsmoker had the odds ratio 0.31 (95% CI: 0.11 - 0.92) times less likely compared with those who were past or current smokers which is statistically significant (p=0.043).

## Discussion

Benign prostate hyperplasia is one of the common conditions that affects older people which constitutes a huge instigation in sexual function, this indicates how imperative it is to investigate the factors linked to erectile dysfunction in patients with BPH in order to develop specific measures and strategies to improve on their way of life specifically the quality of life.

Several efforts have been made to investigate factors that influence sexual function among men which mainly incline in the subjects with diabetes, hypertension, chronic kidney disease, HIV, cancer patients etc. Erectile dysfunction has been found to impinge significantly on the overall quality of life and it leads to stress, depression, marital disharmony, divorce, poor performance at work, and other psychological related problems.

### Characteristics of Respondents

All the participants were males having BPH not on medication with a larger proportion of patients aged from 40 and above. The mean age of respondents was 71 with range of 50 to 98 years. These findings were consistent with the study done by El-Sakka who assessed a total of 374 men with ED using IIEF and the presence of LUTS/BPH, the mean age was 54.8 years (range of 24-84) (5). Therefore, the age of the study population is significant as it portrays the age as one of the factors associated with ED. In addition, care and treatment services will be focused on the age group, i.e., 40 years and above.

### Prevalence of erectile Dysfunction by Age

The study looked at the link between age and LUTS symptoms as well as ED-related disorders. In this study the prevalence of LUTS severity was found to be increasing with age (<61 (55%), 61-70 (82%), and more than 71 (93%). The study showed that the ED increases with age, findings which were consistent with the Classic Massachusetts Male Study which showed at the age of 50, the prevalence of ED is around 50%, rising to 70% at the age of 70.

### Prevalence and severity of erectile dysfunction

About 82% of patients had ED ranging from mild, moderate to severe. These findings were correlating with the study done by Rosen, & Johnson, (8) the results showed the prevalence of ED ranged from 51.8 percent in men with mild LUTS to 80.1 percent in individuals with severe LUTS, according to the study. In both studies, the prevalence of erectile dysfunction is high. This study underlines the need of monitoring sexual functioning in men with BPH and LUTS early in their treatment to ensure that their treatment is as effective as possible. (8)

### Prevalence of erectile dysfunction by Marital status

In this study, it was observed that patients that are married or cohabiting had a higher grade of ED compared to men who were not married with 72.34%, 10.64% and 17.02% respectively. This was similar to another study done by Liang, Guo QingZheng, Jun Biao Wu, found men who aged 40 to 80 years with regular sex partners had erectile dysfunction in 76.18%, (15), this can be explained in the fact that men who are married have responsibilities to provide the family with basic needs so may have stress in terms of socioeconomic status that may have a negative impact to sexual function.

### Prevalence of erectile dysfunction with Smoking and Diabetes Mellitus

In this study cigarette smoking and diabetes mellitus was found to have a prevalence of 56% and 27% respectively. Smoking-related ED is primarily linked to endothelial dysfunction and a decrease in nitric oxide (NO) availability, this was supported by considerable evidence. Cigarette smoking has a variety of negative effects on endothelial cells, including decreased endothelial nitric oxide synthase activity, impaired endothelium-dependent vaso-relaxation, increased expression of cell adhesion molecules and trans-endothelial growth factor, and impaired regulation of key thrombotic factors.

The mechanism that increased advanced glycation end-products (AGEs) and oxygen free radical levels, impaired nitric oxide (NO) synthesis, increased endothelin B receptor binding sites and ultrastructural changes, upregulated RhoA/Rho-kinase pathway, NO-dependent selective nitrergic nerve degeneration, and impaired cyclic guanosine monophosphate (cGMP)-dependent kinase-1 these are all that supports ED in diabetic patients.

## Conclusions

- Erectile dysfunction is associated with lower urinary tract symptoms caused by benign prostatic hyperplasia. The severity of erectile dysfunction is linked to the IPSS, which is defined as symptoms of lower urinary tract severity, and the greater the IPSS, the more severe the symptoms and erectile dysfunction.
- Even though there is a validated tool for assessing the sexual function available and known among health care provider it is not used in routine patients’ consultations.
- Among the factors associated with erectile dysfunction was an increase in age, lower urinary tract symptoms, cigarette smoking, diabetes mellitus and hypertension.

## Recommendations

- This study highlights the importance of enquiring about ED during routine Urology consultations in clinics and wards, using the IIEF score tool at MNH, as well as at all health facilities across the country.
- There is a need of introducing a specialized sexuality clinic as a sub specialty in Urology in order to address the concern of sexual function and be able to manage the problem in order to impact the quality of life in men with BPH.

## Data Availability

The datasets analyzed during this study are available from the corresponding author on reasonable request.

## Abbreviations

BPH: Benign Prostate Hyperplasia
ED: Erectile Dysfunction
IIEF: International Index of Erectile Function
IPPS: International Prostate Symptoms Score
LUTS: Lower Urinary Tract Symptoms
QoL: Quality of Life
MNH: Muhimbili National Hospital
MUHAS: Muhimbili University of Health and Allied Sciences

## DECLARATIONS

### Consent to publication

Not applicable

### Competing interests

The authors declare that they have no competing interests.

### Funding

Tanzania - Ministry of Health and Social Welfare

### Authors’ contributions

Dr. S.A.M contributed to the conception and design of the study, drafted original manuscript and revised the manuscript. Prof.C.M contributed to design of the study, data analysis and also revised the manuscript. Dr.D.M contributed to conception and design of the study, data validation and analysis, and also critically revised the manuscript. Dr.P.M contributed in data acquisition and entry, analyzed and interpreted the data.Prof. O.N contributed to design of the study, review, analysis and also critically revised the manuscript. All authors read and approved the final manuscript.

### Ethical considerations

The study protocol was reviewed and approved by the Institution of Review Board of the Muhimbili University of Health and Allied Sciences (reference number DA.282/298/01.C/) and permission to collect data was obtained from relevant authorities at MNH. All participants were provided a written informed consent prior to participation into the study.

## Acknowledgement

We thank all the staff members of the Urology unit at Muhimbili National Hospital for their tireless support during this study. We also thank all the study participants who have been involved in this research.

## Authors’ information

Not applicable

## Notes

### Competing Interest Statement

The authors have declared no competing interest.

### Funding Statement

This study received fund from Tanzania ministry of health

### Author Declarations

The study protocol was reviewed and approved by the Institution of Review Board of the Muhimbili University of Health and Allied Sciences (reference number DA.282/298/01.C/) and permission to collect data was obtained from relevant authorities at MNH.

